# Development and design specifications for an accelerometer-based biomechanical, prosthetic sensorimotor platform

**DOI:** 10.1101/2020.07.17.20151605

**Authors:** P A Johnson, J C Johnson, A A Mardon

## Abstract

Following stroke, injury, or exposure to physically limiting conditions, limbs can become physiologically compromised. In particular, motor and fine-dexterity tasks involving the arm, particularly in locomotion, grasp and release, can be influenced becoming either delayed and having to deal with greater force demands. Current prosthetic systems use electromyography (EMG)-based techniques for creating functional sensorimotor platforms. However, several limitations in practical use and signal detection have been identified in these systems. Accelerometer-based sensorimotor systems have been suggested to overcome these limitations but only proof-of-concept has been demonstrated. Here, we explore design specifications for accelerometers being developed for prosthetic integration. We have developed optimizations for the current model, evaluated system properties to enhance sensitivity and reduce signal noise, and performed a pilot test using simulation to test this model. The data suggest these novel design parameters can enhance signal detection, when compared to conventional accelerometers. Future avenues should focus on validation of this design prototype in a full prosthetic system.

## 1. Introduction

Currently, approximately 35-40 million people globally require prosthetic or orthotic support after stroke, injury, and/or other conditions affecting limb function [1]. With the growing incidence of musculoskeletal conditions and non-communicable diseases including stroke and diabetes however, this number is expected to double by the middle of this century. Movement and range of motion is significantly reduced and limited for these individuals. As a result, there is an emerging and exponentially increasing need for supporting prosthetic and orthotic support devices.

For affected individuals, performance of motor and fine-dexterity tasks, particularly those involving upper limbs, in locomotion, grasp and release, can be influenced significantly, becoming both delayed and requiring greater force demands. Applied and external acceleration are crucial determinants of these force demands, especially for systems requiring sensory integration and output generation, such as robotic prosthetic limbs. For many of these tasks, appropriate input and output command signals must be determined. Currently, such biomechanical systems have placed a focus on providing both sensory and motor feedback platforms to the user through electromyography (EMG)-controlled prosthesis [2]. EMG utilizes electrical activity between muscle fibres during contraction and relaxation phases to quantify the neuromuscular signalling patterns of the innervating motor units. While it is possible to perform invasive or non-invasive sensing, non-invasive is virtually always preferred, as it would allow surface EMG signal detection from the skin surface alone. However, this technique has several limitations including higher levels of difficulty in identifying signal acquisition sites, high sensitivity for motion and motion artefacts, and overall lower dexterity [2,3]. Compared to this, intramuscular EMG signals using implantable sensors provide enhanced access for signal detection and additionally enable multiple degrees of control to the prosthetic.

Many current prosthetics, including myoelectric prosthetic upper and lower limbs, employ EMG-based methods for sensing, detection, and feedback processing of motor control signals. Over the recent years, there has been a growing body of evidence establishing the utilization of tri-axial MEMs (Micro Electro-Mechanical System) sensor accelerometers as an alternative or composite sensorimotor feedback platform for use in rehabilitation [4–7]. Here, we suggest a novel design for accelerometers for the implementation of this model, which could allow multiple degrees of freedom (DOF) sensors built into prosthetic and orthotic wearable.

## 2. Prosthesis model

Clinical prosthetic require the use of compact elements that include drive systems, cells, and microprocessors for functional motion adjustment and response. Within drive systems, servo motors enable continuous monitoring of feedback signals and on-going adjustment of movement for any deviations detected. Additionally, these system elements must have appropriate interfaces and the ability to sustain constant activity for practical prosthesis. Kyberd and Poulton suggest the use of a tri-axial system whereby sensors and controllers are employed to detect and correct for 1) segment orientation, 2) motion compensation, and 3) inertial platform [4]. This basic model represents an intermediate stage prior to full integration with limbs, where certain actuators and sensors are independently controlled to create a functional system (Figure 1). Segment orientation is a compensatory mechanism for the accelerometer that takes into consideration the gravitational forces and tri-dimensional, spatial alignments in order to accommodate the motor demand accordingly. Motion compensation adapts for the positioning using the surrounding prosthetic limb segment kinematics. Inertial platform uses holistic, mathematical analysis of prosthesis in interaction with an object of interest.

**Figure 1.**
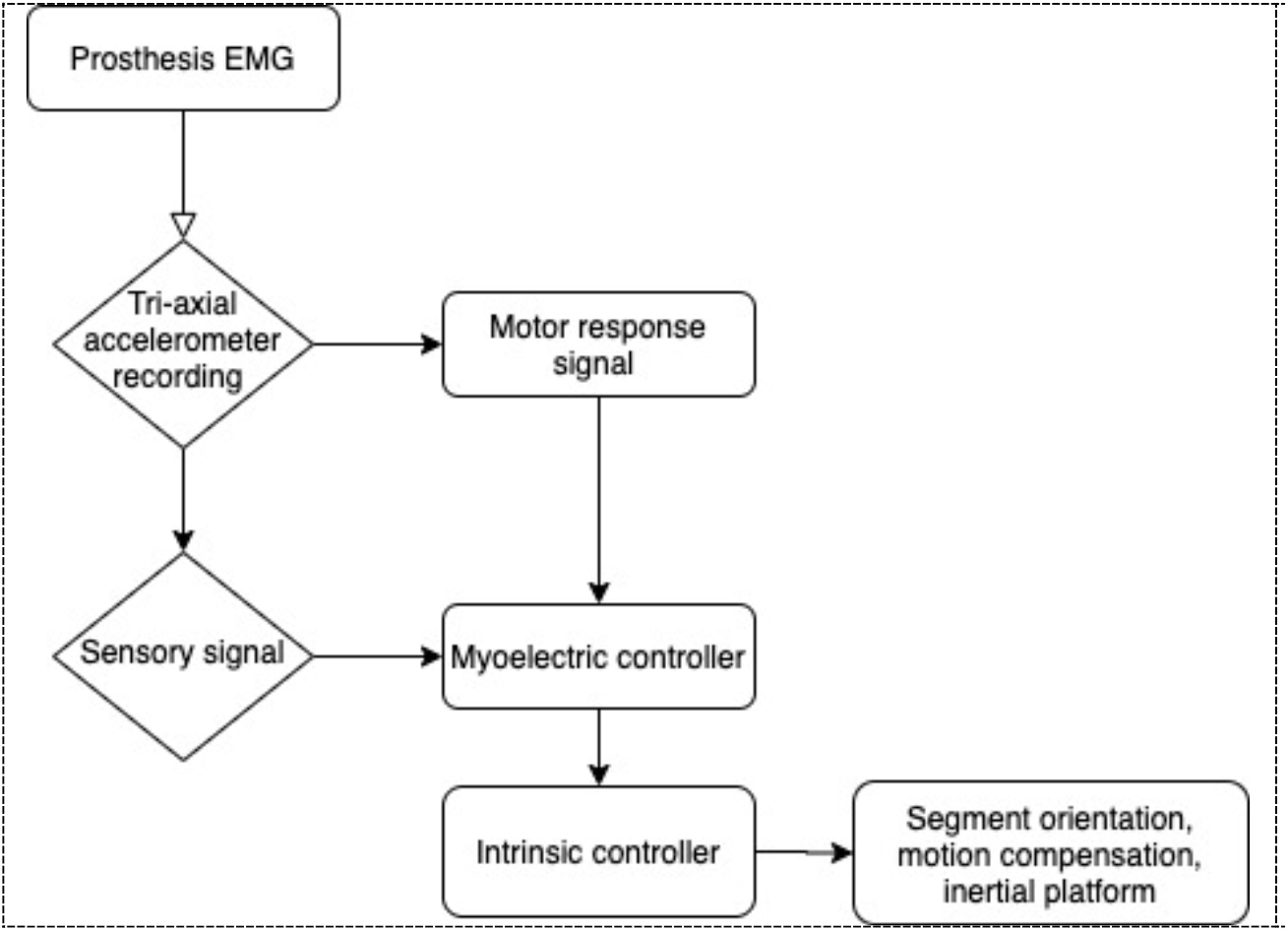
Block diagram of proposed tri-axial model. EMGs are combined with accelerometer sensors to create a functional system that uses sensory and motor command signals to regulate segment orientation, motion compensation, and inertial platform.

While a proof-of-concept has been established for the use of accelerometers, this current model has several limitations, which must first be addressed. First, the measurement of motion requires the input signal to be integrated over time, which results in a high signal noise leading to drift. Although the use of inertial sensors has been described as a technique to reduce this effect, several current models have yet to incorporate these within prosthetic designs, while relying exclusively on EMG-based sensing methods. Additionally, sensitivity, which refers to the ratio of change in output to the change in expected input of the sensor, and the bias, which results from the non-zero of the output signal of acceleration generated by the sensor, are both commonly observed limitations of MEMs sensor accelerometers. Both input and output signals could also be influenced by factors such as material, construction, operation conditions (e.g., temperature, pressure, surrounding electrical and/or mechanical prosthesis elements, etc.) It is additionally possible for factors within a biomechanical system to influence the signal, including levels of strain resulting from applied forces. As accelerometer sensing data also depends significantly on the integration, even a subtle acceleration error could have a large effect on detected data (e.g., in a position estimation test, a double integration of an acceleration error of 0.1*g* can result in a 350 m position error) [8].

Moreover, several proof-of-concept designs limit their models to a single joint with limited range of motion (i.e., transradial prosthetic wrists with only 2 DOFs). In light of the replacement of such joints, there are often times a functional need to establish more advanced ranges of motion. Many studies also make assumptions regarding idealized trajectories, static conditions, downstream output signalling effects, etc. In most cases, it is additionally challenging to analyse real-time data being collected by sensors. The objective of design and development of this novel prototype model was to provide an improved model that addressed many of these inherent limitations.

## 3. Prototype model design and development

Through this prototype model, we made several considerations to update conventional biomechanical prosthetic systems. For considerations at the accelerometer-level, design specifications were developed by our team. In this model, we utilized a template Totally Modular Prosthetic Arm with high Workability (ToMPAW) prosthetic arm design [9]. Figure 2 displays this prosthetic arm design and the site of accelerometer incorporation. Although the accelerometer can be employed at numerous sites on the ToMPAW arm, we selected to examine a focused trans-radial segment capacitating motions in multiple DOFs, namely corresponding to the condyloid (between radius and carpal bones of wrist) and saddle (between trapezium carpal bone and first metacarpal bone) joints.

**Figure 2.**
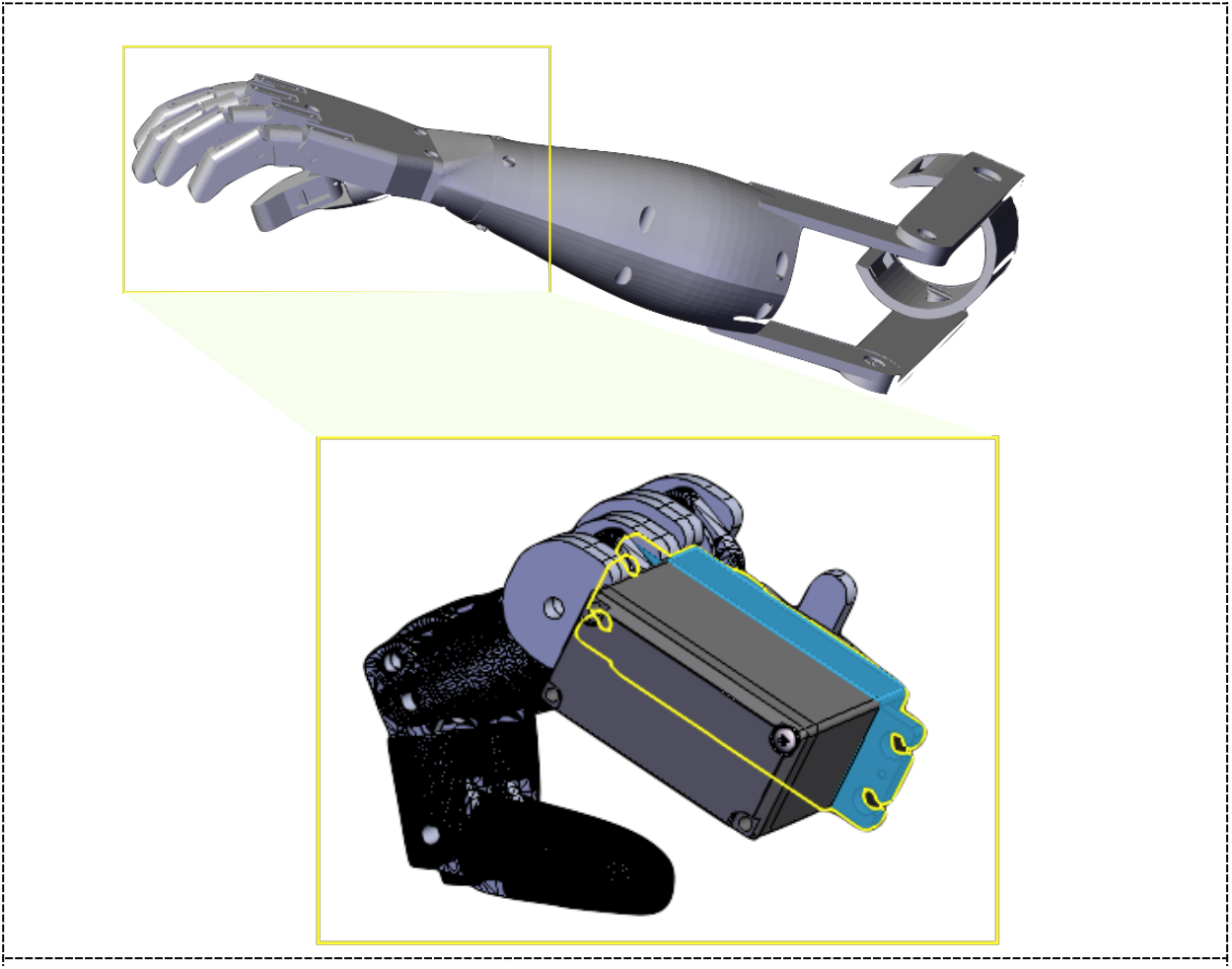
ToMPAW prosthesis arm and accelerometer site.

As the primary aim of this design was to reduce limitations in noise for signal integration, we focused on (i) determining optimal properties for accelerometer and sensor construction and (ii) evaluating its effects on real-time signal quality improvement and feasibility. First, we incorporated support arrays to enable the implementation of additional sensing devices, such as more accelerometers, gyrometers, and magnetometers as required to the conventional design (Figure 3). The most commonly utilized accelerometers include capacitive and piezoelectric accelerometers. Capacitive accelerometers uses a capacitive coupling mechanism and is typically preferred for low-frequency tasks, while piezoelectric accelerometers are transducers that use the accumulated electric charge resulting from applied mechanical stress and typically preferred for high-frequency, vibration and high shock measurements. Both of these types of accelerometers offer a good measure of dynamic changes in acceleration within a system. MEMs-based accelerometers are the conventional standard for accelerometers recommended for prosthetic integration and can use either piezoelectric transduction or capacitive sensing to detect acceleration. While several advantages, such as low temperature sensitivity, linearity, and small footprint, relative to other available techniques, have been suggested for capacitive sensing accelerometers [10], practically, piezoelectric MEMs-based accelerometers have been reported to offer higher sensitivity, good linearity, and lower noise [11]. To enhance current specifications, we decided to utilize piezoelectric MEMs-based accelerometers in our model. For the construction of the accelerometer, we used a standard FR-4 PCB, which consists of woven glass-reinforced epoxy laminate material.

**Figure 3.**
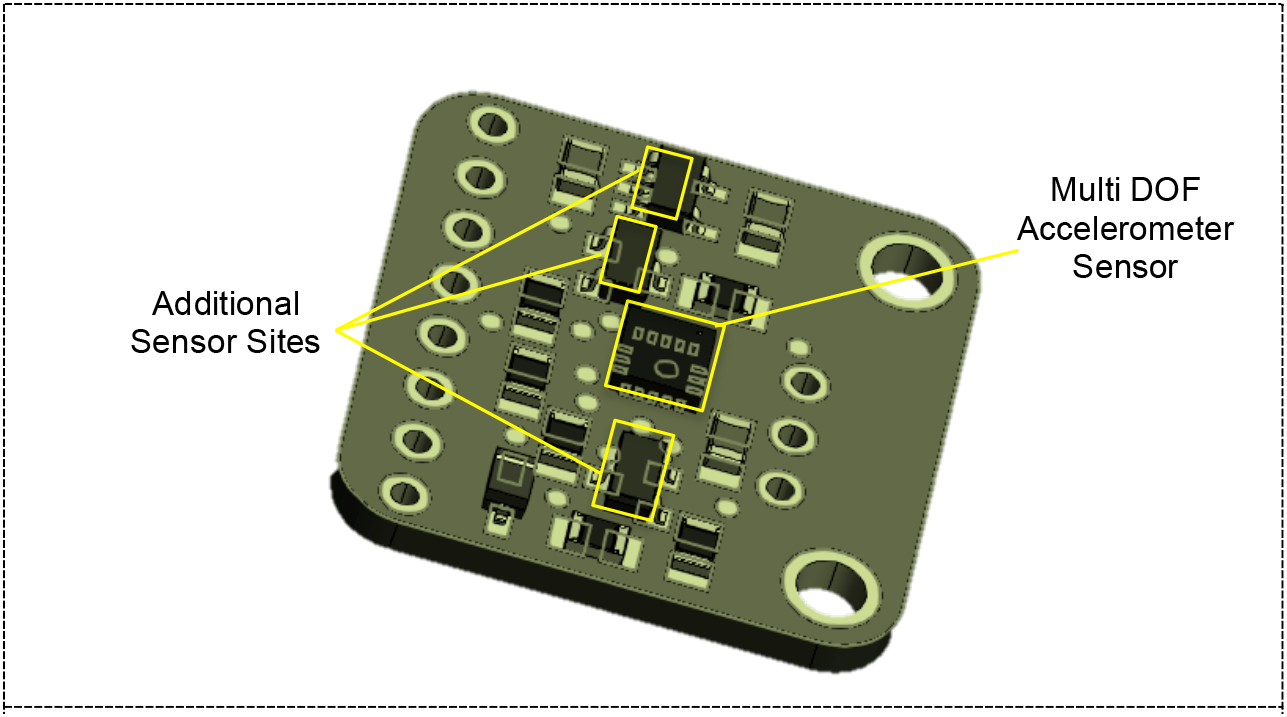
Tri-Axial Accelerometer Prototype Design

While a conventional circuit board unit was utilized, we upgraded the model to incorporate multiple sites for the incorporation of these piezoelectric MEMs-based accelerometer sensors and other elements (e.g., capacitive sensors, gyrometers, magnetometers, thermistor, etc.) We also selected a commercially available, additive piezo-ceramic material for the construction of sensors for long durability and stability, especially under varying temperature ranges, enhancing the sensitivity of current prosthetic accelerometer sensor designs.

In addition to defining specifications for accelerometer construction material, we explored the optimal design considerations for sensor design. A typical MEMS-based piezoelectric accelerometer sensor consists of two basic units, including a cantilever and seismic mass. The physical properties of these sensors determine the sensitivity of measured signals, as well as the level of noise and drift that occurs during movement tasks. In order to account for low sensitivity limitations observed in current models, we designed sensor elements to allow high-resolution measurements. In order to do this, we calculated optimal dimensions, material constants, and sensor properties, including deflection, stress, and strain. Assuming linear constitutive law, the sensor system can be modelled using a mass-spring-damper model where the seismic mass is equivalent to the mass element, *M*, and the cantilever beam is equivalent to the spring and damper elements in a base-excited configuration of the model (Figure 4).

**Figure 4.**
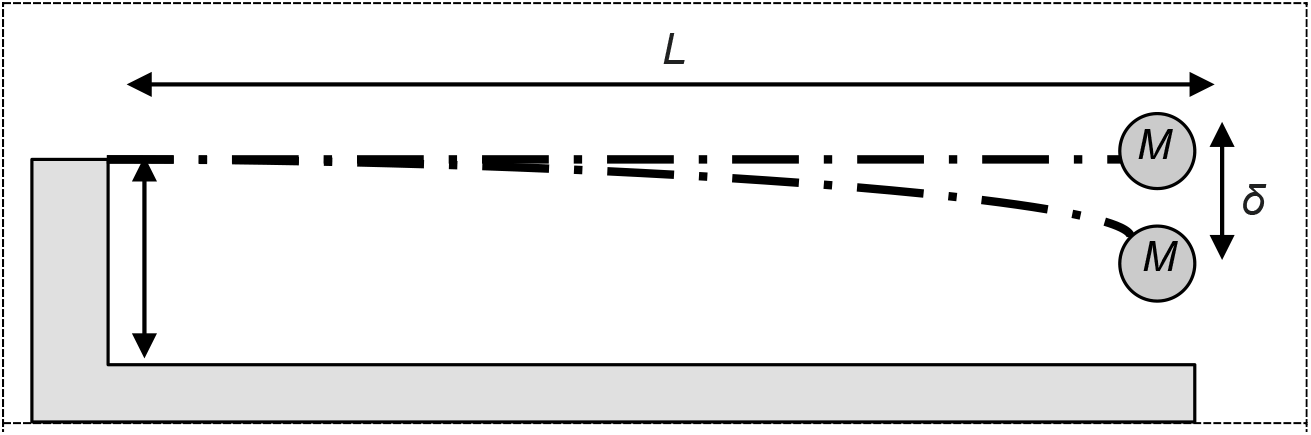
Accelerometer sensor base-excited cantilever model

This model also assumes both the spring and damper are in mechanical parallel and that motor input is received from the base. According to the assumptions made by this model, it is possible to determine optimal design standards for various material properties for sensor construction including:

- *Q-factor*: a measure of how under-damped an oscillator is. As all materials have a natural frequency of vibration, determining the Q-factor can be utilized to measure the dampening according to the construction material properties. If the Q-factor is too low, damping in the system will result in a reduced sensitivity. The Q-factor can be calculated by:

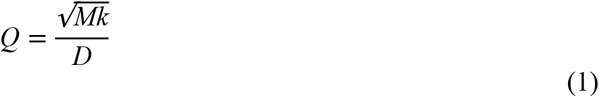

where *Q* is the Q-factor, *M* is inertial mass, *k* is the spring constant, and *D* is the damping coefficient, where

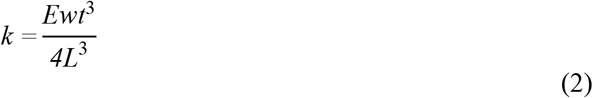

and

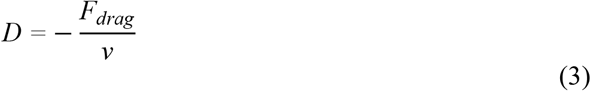

where *E* is Young’s Modulus, *w* is cantilever width, *t* is its thickness, and *L* is its length. In calculation of the damping coefficient, the drag force, *F*_drag_, and velocity, *v*, and ultimately, the rigidity parameter are largely dependent on material properties. By this proof, a sensor design of higher sensitivity must utilize an optimized higher inertial mass, cantilever construction using a material with a higher spring constant.
- *Deflection*: the degree to which a cantilever is displaced under a load. A higher deflection is typically an indicator of a higher cantilever design sensitivity [12]. The Stony Equation can be utilized to calculate deflection:

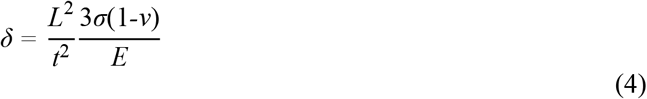

where *δ* is the cantilever deflection, *L* is the length of the cantilever beam, *t* is its thickness, σ represents the uniaxial stress or force per unit square, *v* is Poisson’s ratio, and *E* is Young’s modulus. Deflection is inversely proportional to the spring constant.
- *Stress*: a measure of the internal forces resulting within a material system. Mechanical stress can be classified according to the type of forces acting on the system and include: scalar (tension or compression), bending, and torsional (shear) stress. In a cantilever beam system, the increasing the structural load, *M*, notably increases levels of bending and shearing stress.
- *Strain*: a measure of stretch or deformation that occur as a result of mechanical stress. Strain occurs when a force is applied to an object and results in a physical change, whereas stress concerns only the forces regardless of any physical changes to the material. The resolution of measurements, quantifiable by the Q-factor, and deflection levels, rely essentially on both of these parameters. This is demonstrated mathematically by the Young’s modulus ratio and its presence as a variable in calculations for the spring constant (ultimately, Q-factor) and deflection:

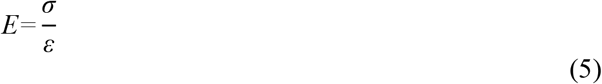

where *σ* is the uniaxial stress, determined as a pressure or force per square unit, and *ε* is the strain, which is a dimensionless quantity representing the proportion of deformation. Based on optimization used in the design of multi-DOF robotic arms in varying gravitational fields to enhance load bearing [13], it is possible to describe optimal vibration frequencies and strain magnitudes. Both stress and strain therefore influence the overall resolution of measurements and must be considered in the design.

In Table 1, we summarize the changes that are required according to these design parameters in order to enhance the quality of measurement signals.

**Table 1.** Design parameter optimization to enhance signal detection quality.

| Parameter | Description |
| --- | --- |
| Q-factor | Low Q-factors results in damping, which achieves a lower sensitivity within a system. As such, a system with higher Q-factors is favoured in an optimization. |
| Inertial mass | Higher inertial masses are preferred to increase system sensitivity. However, masses must be optimized to ensure the structural stability of the system. |
| Spring constant | Materials with higher spring constants (e.g., piezo-ceramics), which also tend to have associated modulus parameters, favour higher sensitivity systems. |
| Rigidity | Within a system, the sensor's damping coefficient, which is based on the material's resistance to the drag force at a given velocity, should be low to ensure a higher Q-factor. |
| Cantilever dimensions | Length, thickness, and width of the cantilever affect vibrational frequency capacities of the sensor. Higher design sensitivity features an inverse relationship between length and thickness producing higher deflection and Q-factor values. Width also influences the spring constant of the system. |
| Deflection | Higher deflection is an indicator for a higher cantilever design sensitivity and associated with lower signal noise events |
| Young's Modulus | A material's resistance to applied mechanical stresses and strain capacity are key variables in deflection and Q-factor measurements in the system. Typically, a higher strain capacity and lower stresses are achieved in an ideal system. |

## 4. Design Specifications

Based on these properties, design specifications were developed for a novel prototype using fixed accelerometer dimensions. In Table 2, we report the above optimization equations, modelling, and feasibility parameters, which approximated threshold specifications for novel accelerometer design. In Table 3, we compare technical specifications between existing conventional and MEMs accelerometers and the novel design prototype.

**Table 2.** Threshold specifications for accelerometer parameters calculated according to optimization equations and earlier models for novel design prototype.

| Parameter | Specification |
| --- | --- |
| Accelerometer Mass (Fixed) | 150 $\mu\text{g}$ |
| Accelerometer Dimensions (Fixed) | 750 $\mu\text{m} \times 250 \mu\text{m}$ |
| Minimum Cantilever Length | 210 $\mu\text{m}$ |
| Maximum Cantilever Thickness | 18 $\mu\text{m}$ |
| Vibration Resonance Frequency | 120 Hz |
| Average Acceleration | 2.5 m/s/s |
| Piezoelectric Sensor Material | modified $\text{PbTiO}_3$ |

**Table 3.**
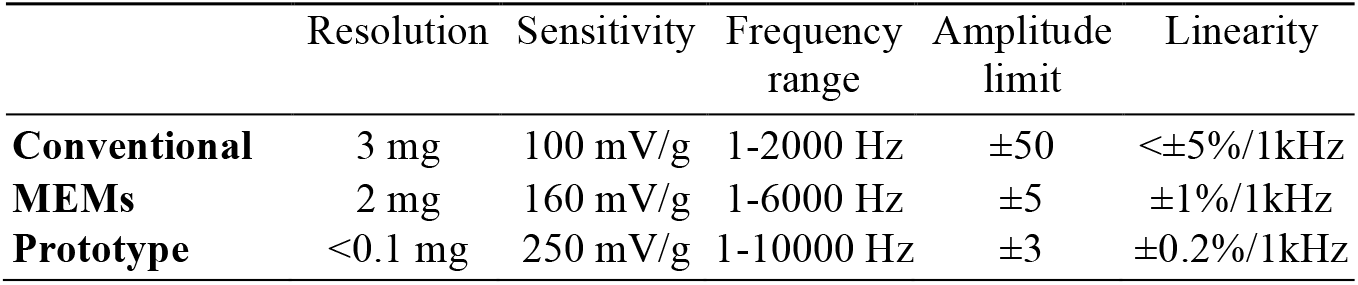
Comparison of novel design prototype to existing accelerometer designs.

## 5. Simulation Modelling

To test optimizations, we employed MATLAB to perform simulations using pre-existing prosthetic movement acceleration dynamics data. Modelling was based on crank-slider mechanism optimization, which uses predictive and output data to calculate position, crank velocity, and acceleration over time [14]. We utilized available tracked prosthetic movement data with six DOFs and generated acceleration over time plots to compare the signal-to-noise ratio and drift between conventional accelerometers and our novel design. Figure 5 displays the dynamics data capturing translational movement along an X-axis, translation along a Y-axis, translation along a Z-axis, rotation around a roll axis, rotation around a pitch axis, and rotation around a yaw axis. This data includes movement requiring segment orientation, motion compensation and an inertial platform. In Figure 6, results of the simulations for a calibrated conventional accelerometer model and our novel prototype design model are shown. It displays the difference in signal quality during segment orientation, motion compensation, and inertial platform. As a result, these algorithms can then used to generate command outputs in a prosthetic system. Of the two, our prototype was determined to reduce signal-noise effects observed in conventional accelerometers. Our modelling and prototype therefore demonstrates it is not only possible to mechanically dampen a system, but also that we can reduce noise and increase signal sensitivity by upgrading accelerometer design specifications.

**Figure 5.**
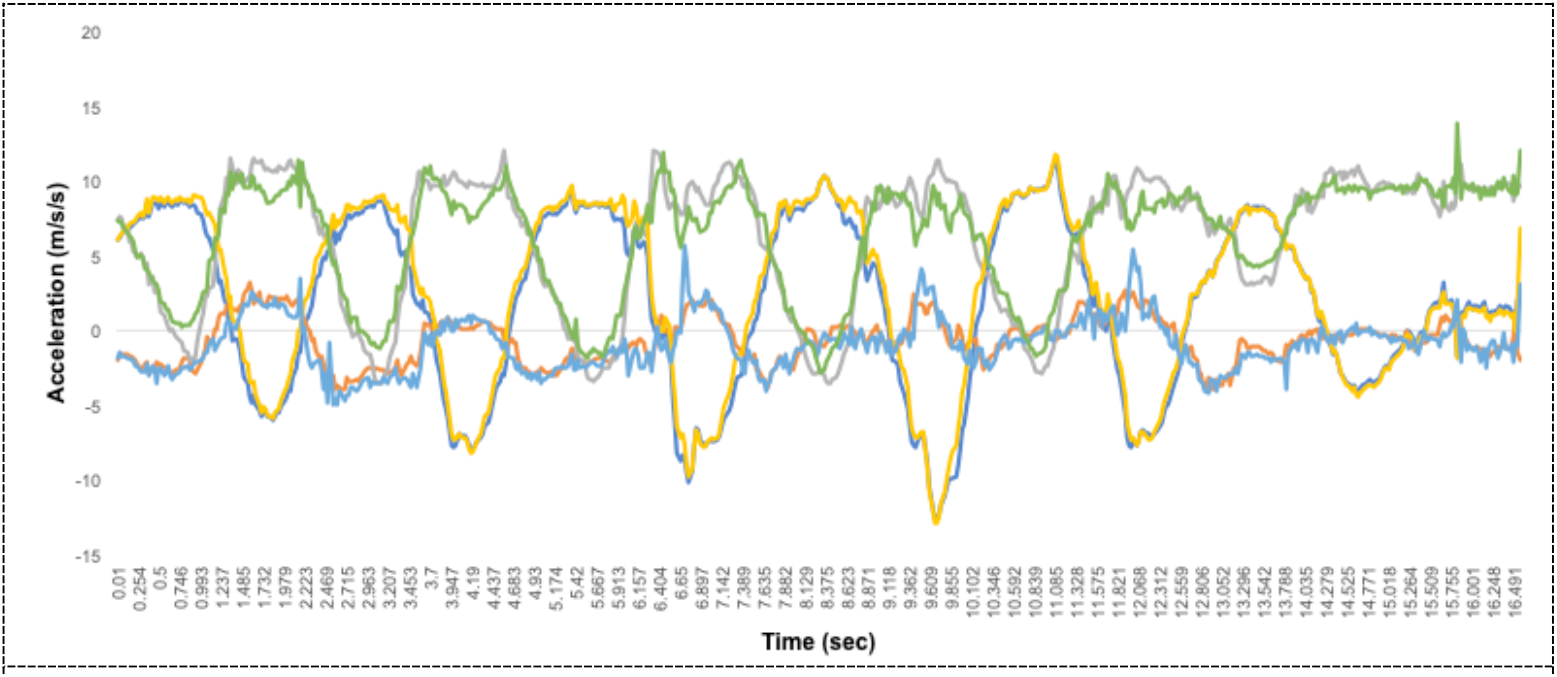
Raw tracked prosthetic movement acceleration dynamics data.

**Figure 6.**
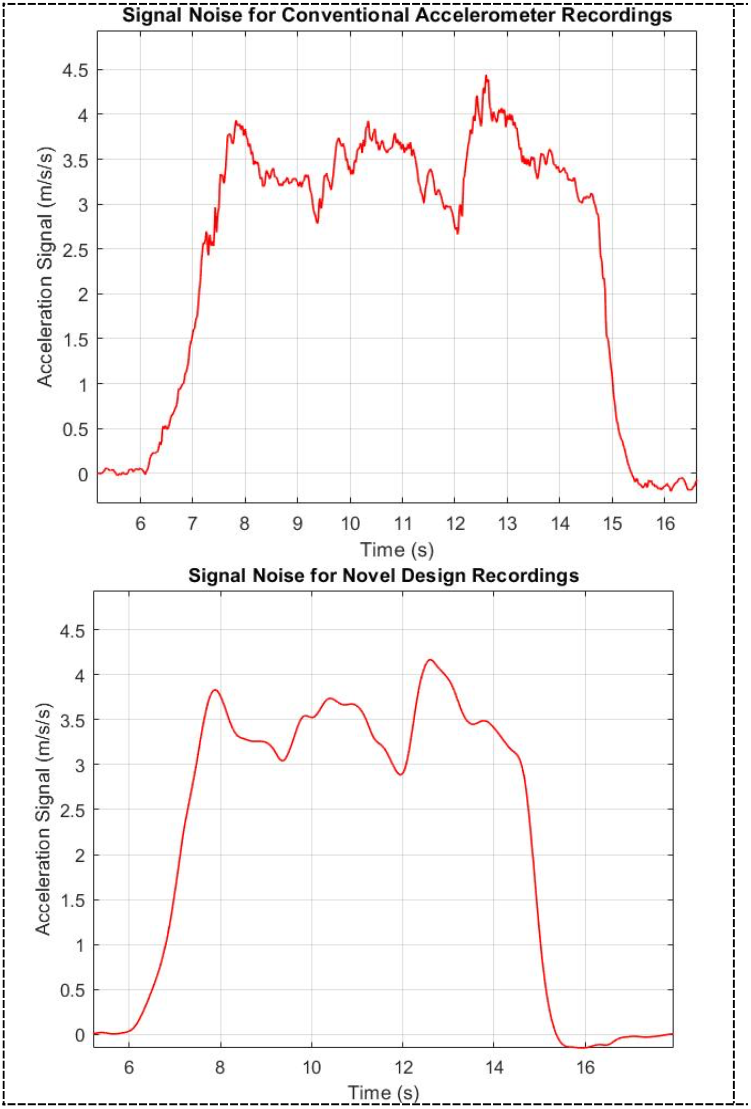
Signal-noise reduction in novel design prototype. Using acceleration dynamics data for prosthetic arm movement using six degrees of freedom, an integrated acceleration value was generated using a model on MATLAB for a conventional accelerometer sensor design and the design for the novel accelerometer prototype.

## 6. Limitations and Future Avenues

While promising, this proposed model still has several limitations and integral design considerations must be made before its implementation. First, it is a proof-of-concept having been tested only via simulation and must be evaluated in a prosthetic system. Our simulation also focuses on two main accelerometer subtypes in modelling signal quality and would benefit from examination of other subtypes. It is additionally necessary to validate these results by a comparative analysis with concurrent EMG measurements to ensure the recording is indeed detecting a motor signal within the prosthetic system. While this novel prototype could overcome several limitations with EMG sensorimotor systems in limb prosthesis, extensive work must be done to model movements using six or more DOFs. Thus, future avenues should also focus on exploring multi-DOF sensor application, which may significantly reduce the time and cost required to perform manoeuvres. Another of area of interest should focus on real-time noise filters and integrating previously described strain devices for osseo-integrated prosthesis devices, which can be optimized for bone adaptation.

## 7. Conclusion

We have proposed a model for optimization, system properties and considerations for its implementation, described a novel piezoelectric MEMS accelerometer prototype design for prosthetic integration, and demonstrated a proof-of-concept using a simulation to show enhanced signal quality detection. Future studies should focus on incorporation of this design and real world testing in complete prosthesis systems.

## Data Availability

All data supporting the findings of this study are available from the corresponding author [PAJ] on request.

## References

[1] World Health Organization 2017 Standards for Prosthetics and Orthotics - Part 1: Standards (Geneva: World Health Organization)

[2] Sudarsan L S and Sekaran E C 2012 Design and development of EMG controlled prosthetics limb Procedia Engineering vol 38 (Elsevier Ltd) pp 3547–51

[3] Geethanjali P 2016 Myoelectric control of prosthetic hands: State-of-the-art review Med. Devices Evid. Res. 9 247–55

[4] Kyberd P J and Poulton A 2017 Use of Accelerometers in the Control of Practical Prosthetic Arms IEEE Trans. Neural Syst. Rehabil. Eng. 25 1884–91

[5] Yang J, Kusche R, Ryschka M and Xia C 2018 Wrist movement detection for prosthesis control using surface EMG and triaxial accelerometer Proceedings - 2017 10th International Congress on Image and Signal Processing, BioMedical Engineering and Informatics, CISP-BMEI 2017 vol 2018-January (Institute of Electrical and Electronics Engineers Inc.) pp 1–6

[6] Culhane K M, O’Connor M, Lyons D and Olaighin G 2005 Accelerometers in rehabilitation medicine for older adults Age Ageing 34 556–60

[7] Klisic D, Kostic M, Dosen S and Popovic D 2009 Control of prehension for the transradial prosthesis: Natural-like image recognition system J. Autom. Control 19 27–31

[8] Kowalczuk Z and Merta T 2014 Modelling an accelerometer for robot position estimation 2014 19th International Conference on Methods and Models in Automation and Robotics, MMAR 2014 (Institute of Electrical and Electronics Engineers Inc.) pp 909–14

[9] Kyberd P J, Poulton A S, Sandsjö L, Jönsson S, Jones B and Gow D 2007 The ToMPAW modular prosthesis: A platform for research in upper-limb prosthetics J. Prosthetics Orthot. 19 15–21

[10] Mohammed Z, Elfadel I and Rasras M 2018 Monolithic Multi Degree of Freedom (MDoF) Capacitive MEMS Accelerometers Micromachines 9

[11] Albarbar A, Mekid S, Starr A and Pietruszkiewicz R 2008 Suitability of MEMS accelerometers for condition monitoring: An experimental study Sensors 8 784–99

[12] Ansari M Z and Cho C 2009 Deflection, frequency, and stress characteristics of rectangular, triangular, and step profile microcantilevers for biosensors Sensors 9 6046–57

[13] Johnson J C, Johnson P A and Mardon A A 2020 Development of a Mechanical Strain Device to Prevent Microgravity-Induced Bone Loss Pac. J. Sci. Technol. 21 274–5

[14] Halicioglu R, Dulger L C and Bozdana A T 2014 Modelling and Simulation Based on Matlab/Simulink: A Press Mechanism Related content Modelling and Simulation Based on Matlab/Simulink: A Press Mechanism J. Phys. Conf. Ser. 490 012053

